# Where risk becomes visible: a layered fixed-policy framework for diabetic kidney disease screening in type 2 diabetes

**DOI:** 10.64898/2026.04.21.26351384

**Authors:** Ahmed Khattab, Zhe Wang, Vinodh Srinivasasainagendra, Hemant K. Tiwari, Ruth Loos, Nita Limdi, Marguerite Ryan Irvin

## Abstract

**Background:** Diabetic kidney disease (DKD) is a leading cause of kidney failure in individuals with type 2 diabetes (T2D), yet risk identification in routine clinical practice remains incomplete. A critical and often overlooked barrier is risk observability: how much of a patient’s underlying risk is actually captured in their clinical record at the time of screening. Existing prediction models evaluate performance using model-specific thresholds, making it difficult to understand how additional data sources alter real-world screening behavior or which individuals benefit when models are expanded.

**Methods:** We developed a series of five nested machine learning models evaluated at a one-year landmark following T2D diagnosis using data from the All of Us Research Program (N = 39,431; cases = 16,193). Each successive model added a distinct information layer -- intrinsic risk, laboratory snapshots, medication exposure, longitudinal care trajectories, and social determinants of health (SDOH) -- while retaining all prior features. All models were evaluated under a fixed screening policy targeting 90% specificity, so that the false positive rate remained constant as the information available to the model grew. External validation was conducted in the Bio*Me* Biobank (N = 9,818) without retraining.

**Results:** Discrimination improved consistently across layers, from AUROC 0.673 (M1) to 0.797 (M5). Under the fixed screening policy, sensitivity nearly doubled from 0.27 to 0.49, with a cumulative recovery of 30.4% of cases missed by the base model. Gains were driven by distinct subgroups at each transition: laboratory features identified biologically high-risk individuals; medication features captured those with high treatment intensity reflecting advanced cardiometabolic burden; longitudinal care trajectory features rescued cases with biological instability observable only through repeated measurements; and SDOH features recovered individuals with limited clinical observability, with rescue probability highest among those with the fewest recorded monitoring domains. Sparse data in the clinical record indicated low observability, not low risk. Social and genetic features each contributed most when downstream physiologic signal was limited, supporting a contextual rather than universal role for each. In Bio*Me*, discrimination was attenuated (M4 AUROC 0.659), but the relative ordering of information layers was fully preserved, and a systematic upward shift in predicted probability distributions underscored the need for recalibration before deployment in a new setting.

**Conclusions:** DKD risk detection in T2D is substantially improved by integrating complementary information layers under a fixed clinical screening policy, with gains arising from distinct domains that identify at-risk individuals in different clinical contexts. The layered landmark framework introduced here reveals how risk observability -- shaped by monitoring intensity, healthcare engagement, and access -- determines what a screening model can detect, and provides a foundation for context-aware EHR-based screening that accounts for data availability at the time of risk assessment.

Graphical abstract.
Study design and layered DKD screening framework
The top row defines the cohort timeline, in which predictors are derived from clinical data collected between T2D diagnosis and the 1-year landmark, and incident DKD is ascertained after the landmark. The second row depicts the nested model architecture, in which five successive models sequentially incorporate intrinsic risk, laboratory snapshot features, medication exposure, longitudinal care trajectories, and social determinants of health, while retaining all features from prior layers. The third row summarizes model development in the All of Us Research Program (N = 39,431) and external validation in the Bio*Me* Biobank (N = 9,818), where the same trained models and risk thresholds were applied without retraining. The bottom row highlights the three evaluation domains: predictive performance, fixed-policy screening, and missed-case recovery context.
DKD, diabetic kidney disease; T2D, type 2 diabetes; PRS, polygenic risk scores; AUROC, area under the receiver operating characteristic curve; AUPRC, area under the precision-recall curve; PPV, positive predictive value; SHAP, SHapley Additive exPlanations.

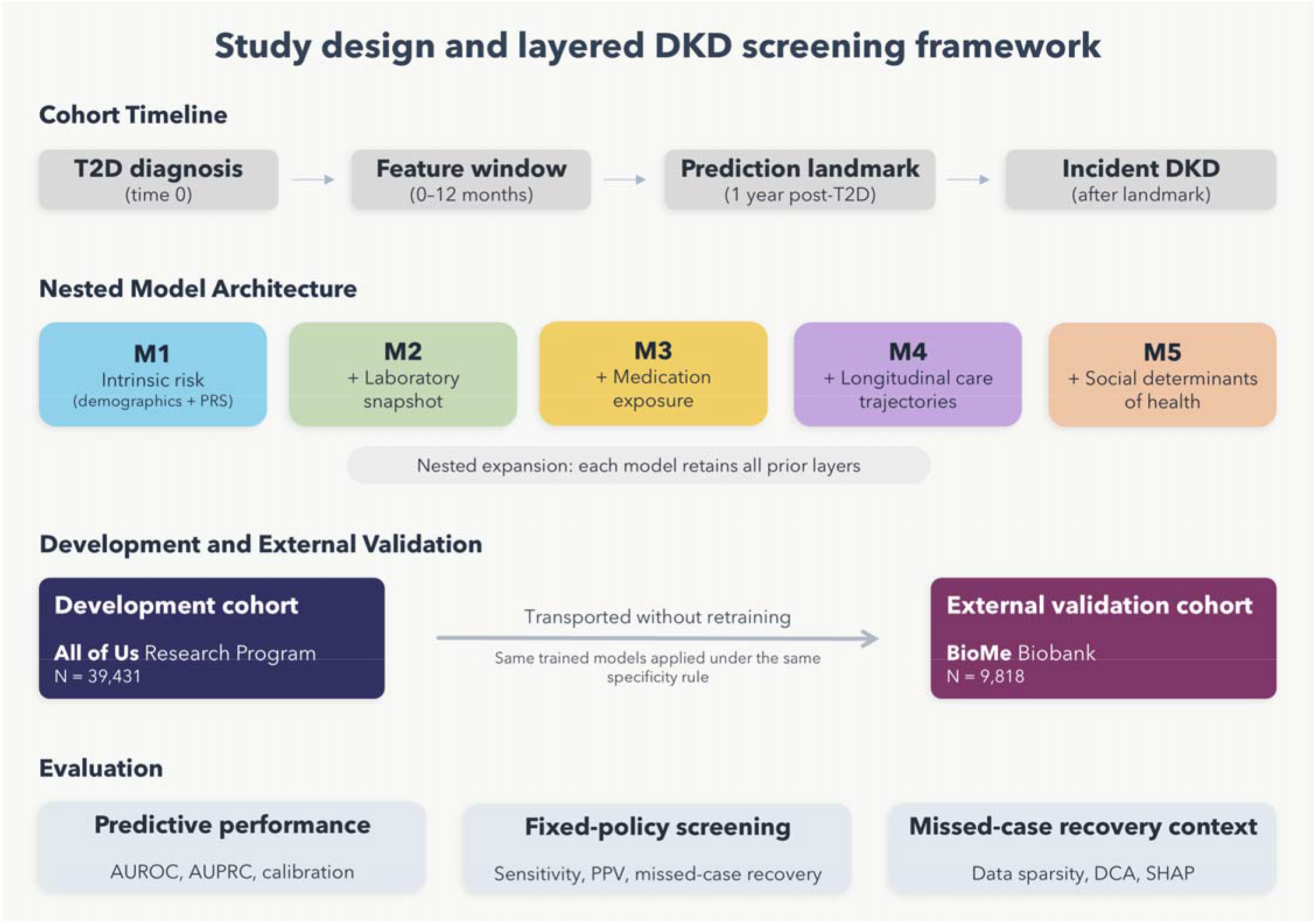

## Introduction

Diabetic kidney disease (DKD) is one of the most common and serious complications of type 2 diabetes (T2D). It contributes substantially to morbidity, mortality, and healthcare costs worldwide.^1^ A substantial proportion of individuals with T2D develop DKD during their lifetime, and it remains a leading cause of chronic kidney disease (CKD) and kidney failure globally. Clinical guidelines emphasize early identification to enable timely intervention, yet DKD is frequently detected only after substantial and potentially irreversible injury has occurred.^2–5^

Despite its clinical importance, early identification of individuals at risk for incident DKD remains difficult in routine clinical practice. Risk does not emerge in the same way for all patients. The transition from T2D to DKD is shaped by biology, metabolic state, treatment, and patterns of clinical care. It is also shaped by what information is captured in the clinical record. Two individuals with similar underlying risk may differ in when and how that risk becomes visible in the electronic health record (EHR). These differences may reflect variation in monitoring intensity, healthcare engagement, and access to care.^3,6–8^ We refer to this as risk observability: how much of a patient’s underlying risk is actually captured in their clinical record at the time of screening.

Most current approaches to DKD risk prediction rely on isolated laboratory values, diagnosis codes, or limited sets of clinical risk factors.^9^ These features are informative, but they provide only a partial view of risk before DKD onset. In particular, the absence of an abnormal laboratory value may reflect lack of measurement rather than absence of risk. Snapshot-based models cannot fully address this distinction. Longitudinal changes in kidney and glycemic markers, treatment intensity, and variation in healthcare engagement all influence what information is available when risk is assessed.^3,10,11^

Recent machine learning (ML) studies have shown that DKD risk prediction can improve when more diverse EHR data are included.^12–14^ However, these models are usually compared using global performance metrics and different decision thresholds for classifying patients as high risk, making it difficult to determine how added data sources change case detection in practice. In clinical settings, screening programs operate under a consistent false positive constraint -- deciding how many low-risk patients they are willing to flag incorrectly. When models are compared without holding this constraint constant, it becomes impossible to isolate whether richer data actually changed which patients were identified. Moreover, these models often overlook the role of care patterns and social context in shaping whether underlying risk can be detected from the clinical record.^15^

Social determinants of health (SDOH), including insurance status and social support, can influence patterns of care and follow-up. They may therefore affect how disease risk becomes visible in the clinical record even before overt pathology is documented.^15,16^ However, the value of incorporating SDOH into DKD screening models is unlikely to be uniform across patients. SDOH may be most informative when biological and longitudinal care data are sparse, but this has not been directly tested within a unified, layered prediction framework.

Genetic susceptibility represents another dimension of risk that exists before clinical disease becomes apparent.^17,18^ Polygenic risk scores (PRS) capture inherited predisposition and have been associated with kidney outcomes in population studies.^19–21^ However, their role in DKD screening remains uncertain, and they may be most useful as a complement to clinical data rather than as a standalone predictor. How PRS interacts with longitudinal clinical data, monitoring patterns, and social context -- and whether genetic risk adds value beyond what is already observable in the clinical record -- has not been well characterized.

In this study, we frame DKD risk prediction as a problem of risk observability rather than discrimination alone. Using routinely collected EHR data from a large and diverse United States cohort, we developed a series of nested ML models at a one-year landmark after T2D diagnosis, using only information available before risk assessment. Critically, all models were evaluated under a fixed screening policy -- a consistent specificity target of 90% was applied across all models, so that the false positive rate remained constant as the information available to the model grew. This allowed us to ask directly: as richer data are added, who gets detected that was not identified before? The models sequentially incorporated intrinsic risk, laboratory snapshots, treatment exposure, longitudinal care trajectories, and SDOH. This framework allowed us to test how successive information layers changed detection of incident DKD on a comparable screening basis. We then evaluated its generalizability in an independent EHR-linked biobank with different population characteristics and care patterns.

## Methods

### Data source and study population

We conducted a retrospective cohort study using data from the All of Us Research Program (AoU), a large and diverse United States research cohort that integrates EHR, survey responses, and genomic data.^22^ Participants included adults with established T2D and available longitudinal clinical data. External validation was conducted in the Bio*Me* Biobank, an independent EHR-linked biobank consisting of patients under care in the Mount Sinai healthcare system with a diverse demographic composition reflective of New York City, NY.^23^

Participants were followed longitudinally using routinely collected EHR data. We defined a fixed clinical assessment point at one year following T2D diagnosis, referred to throughout as the landmark date. Clinical features, laboratory measurements, medication exposures, and SDOH were derived from records occurring before the landmark date, as described below.

### Type 2 diabetes ascertainment

T2D was defined using a harmonized phenotype incorporating both laboratory evidence and diagnosis codes, consistent with established clinical criteria. Individuals were classified as having T2D if they met one or more laboratory-based criteria, including hemoglobin A1c (HbA1c) of at least 6.5%, fasting plasma glucose (FPG) of at least 126 milligrams per deciliter (mg/dl), or random plasma glucose (RBG) of at least 200 milligrams per deciliter (mg/dl).

Diagnosis codes from the International Classification of Diseases, Ninth Revision (ICD-9) and Tenth Revision (ICD-10) were used to support ascertainment but were not sufficient in isolation. Laboratory-based criteria were required to occur prior to or concurrent with diagnosis codes to reduce misclassification arising from rule-out diagnoses or coding errors. The specific ICD-9 and ICD-10 codes used to identify T2D cases are detailed in Supplementary Table S1.

When multiple qualifying laboratory measurements or diagnosis codes were present, the earliest qualifying date was assigned as the T2D diagnosis date and used as the T2D index date for cohort entry. Individuals with evidence of type 1 diabetes or gestational diabetes were excluded.

### Diabetic kidney disease ascertainment

Incident DKD was defined using a composite phenotype based on persistent abnormalities in kidney function or albuminuria occurring after the onset of T2D. DKD events were identified using a combination of laboratory criteria and diagnosis codes consistent with established clinical definitions.

Laboratory based DKD criteria included either a reduced estimated glomerular filtration rate below 60 milliliters per minute per 1.73 square meters (eGFR; mL/min/1.73 m^2^) or an elevated urinary albumin to creatinine ratio (UACR) of at least 30 milligrams per gram (mg/g). To ensure chronicity and avoid capturing transient abnormalities, laboratory criteria were required to be persistent, defined as at least two abnormal measurements separated by a minimum of 90 days.

When eGFR was not directly reported, it was calculated from serum creatinine using the CKD Epidemiology Collaboration (CKD EPI) equation. UACR was calculated from urine albumin and urine creatinine measurements when these values were reported separately.

Diagnosis codes from ICD-9 and ICD-10 indicating CKD attributable to diabetes were used to support DKD ascertainment but were not sufficient in isolation. Procedure codes from the Current Procedural Terminology (CPT) system indicating dialysis or kidney transplantation were used to identify and exclude individuals with evidence of advanced kidney disease or renal replacement therapy prior to diabetes onset. The specific ICD-9 and ICD-10 codes used to identify DKD cases are detailed in Supplementary Table S1. Incident DKD was defined as the earliest date at which persistent laboratory abnormalities or qualifying clinical diagnoses were first observed following the T2D index date.

### Landmark prediction framework

Risk prediction was performed using a landmark framework in which all predictors were restricted to information recorded before a fixed clinical assessment point, and the outcome was defined as incident DKD occurring after that point. The primary analysis was conducted at the one-year landmark, one year following the T2D index date, selected to balance two competing considerations. First, a full year of follow-up allows meaningful accumulation of longitudinal clinical data: repeated laboratory measurements, evolving treatment patterns, and early trajectories of kidney and glycemic markers that are not yet available at or shortly after T2D diagnosis. Second, because the majority of incident DKD in this cohort occurred after two years from T2D diagnosis (median time to DKD: 4.05 years; 74% of cases occurring after 2 years), a one-year prediction window preserves a clinically meaningful intervention opportunity for most at-risk individuals.

At the one-year landmark, the risk set comprised individuals who had not developed DKD before the landmark date, who remained under EHR observation at that time, and for whom at least one year of follow-up was available following T2D diagnosis. Individuals who developed DKD prior to the landmark were excluded from the primary analysis (n = 1,661; approximately 10% of incident cases). Predictors were constructed exclusively from clinical information recorded before the landmark date, ensuring strict temporal separation between predictors and outcomes.

To characterize how risk detectability changes as clinical information accumulates over time, secondary analyses were conducted at three additional landmarks: 30 days, 6 months, and 2 years following T2D diagnosis. Results for these analyses are reported in the Supplement.

### Modeling framework

To disentangle the contributions of fixed or intrinsic patient characteristics, cross-sectional laboratory measurements, treatment exposure, longitudinal care patterns, and social context to DKD risk detection, we constructed a series of nested prediction models evaluated at the one-year landmark. Each successive model added a distinct layer of information while retaining all features from preceding models. This design enabled paired, patient-level evaluation of how additional information altered risk classification under a fixed clinical screening policy, rather than comparing independently optimized models.

All models were trained using the same individuals and identical data splits, ensuring that observed performance differences reflected the incremental contribution of added information layers rather than changes in cohort composition, outcome prevalence, or sampling variability. The same nested structure was applied at secondary landmarks to enable direct comparison across prediction timepoints.

For clarity, the nested models are referred to as M1 through M5 throughout the manuscript and figures. M1 included intrinsic risk features only; M2 added laboratory snapshot features; M3 further incorporated medication exposure features; M4 added longitudinal trajectory features; and M5 additionally incorporated SDOH. The features incorporated in each stage of the modeling process (M1-M5) are detailed in Supplementary Table S2.

### Feature construction

#### Intrinsic risk features

The base model incorporated intrinsic risk features expected to be relatively stable over time and largely independent of healthcare utilization intensity. These included age, sex, comorbid conditions, family history of diabetes or kidney disease, and PRS. Comorbidities were defined using diagnosis codes recorded prior to the landmark date to avoid leakage of post-outcome information.

### Laboratory snapshot features

Laboratory snapshot features reflected the patient’s biological state at the landmark date and consisted of the most recent available values recorded before the landmark date for glycemic, renal, metabolic, cardiovascular, hepatic, and hematologic biomarkers.

### Medication exposure features

Medication features captured exposure to major therapeutic classes relevant to diabetes management, hypertension, cardiovascular disease, and kidney protection. All medication features were restricted to prescriptions recorded before the landmark date. For each medication class, we included indicators of recent exposure in the 30 days before the landmark and of any prior exposure before the landmark. Together, these features captured current and cumulative treatment history.

### Longitudinal trajectory features

These features summarize longitudinal trajectories of laboratory values and vital signs to capture both biological evolution and patterns of clinical monitoring over time. For each laboratory test and vital sign, all measurements recorded prior to the landmark date were included, with strict exclusion of measurements occurring on or after the landmark date to prevent information leakage.

Longitudinal features captured complementary aspects of each patient’s clinical course, including summary measures of central tendency and variability, directional trends over time, and recency of measurement. Slopes were estimated at the individual level using ordinary least squares regression of laboratory values on time (years since first measurement), yielding units per year. Slopes were computed only when at least three measurements were available; otherwise, slope features were set to a missing value. Recency features were defined as the elapsed time between the most recent measurement and the landmark date, capturing the intensity and continuity of longitudinal monitoring within each clinical domain.

In addition, clinically interpretable abnormality features were derived for selected laboratory tests, including indicators of whether values crossed clinically relevant thresholds and the proportion of measurements that were abnormal over time. Together, these trajectory-based features encode both biological instability and clinical observability, distinguishing individuals with sparse or infrequent data from those with actively monitored and evolving disease patterns.

At earlier landmarks (30 days and 6 months), trajectory features were excluded and only cross-sectional summary features were retained, as the limited observation window does not support reliable trajectory estimation. This landmark-adaptive feature strategy ensures that models at each time point use only the feature types that are statistically and clinically meaningful given the available data.

### Social determinants of health (SDOH)

SDOH included insurance status and type, employment and marital status, and psychosocial measures such as stress, social isolation, and mistrust. These variables were derived from standardized survey instruments administered as part of the AoU Research Program. Social features were incorporated to capture contextual and access-related factors that may influence both disease risk and the visibility of that risk within the healthcare system.

### Polygenic risk score construction

PRS were computed using AoUPRS, a scalable framework for polygenic score calculation designed for the AoU program.^24^ Scores were derived using published GWAS summary statistics for kidney-related and cardiometabolic traits. Weight tables from the PGS Catalog^25^ were used to compute PRS from whole-genome sequencing data in AoU and from GSA/GDA genotyping array data imputed to the TOPMed reference panel (version 2) in Bio*Me*. PRS were included as intrinsic risk features across all models and landmarks. Trait descriptions and corresponding PGS IDs are provided in Supplementary Table S3.

### Missing data handling

Missing laboratory and longitudinal values were handled natively by the gradient-boosted decision tree algorithm, which learns how to route missing values at each decision point without requiring imputation. For survey-derived SDOH variables, explicit missingness indicators were included to distinguish absence of response from absence of risk. Measurement sparsity was partially encoded through recency and count features, allowing the model to differentiate low monitoring intensity from biologically stable values. No outcome information was used to impute predictors.

### Model training and calibration

All models were implemented using gradient-boosted decision trees (XGBoost).^26^ Hyperparameters were optimized within the training set using Bayesian optimization (Optuna)^27^ with five-fold stratified cross-validation to maximize discrimination. The dataset was split once into training, calibration, and test sets in a 60%, 20%, and 20% ratio and reused across all models within each landmark cohort. The same split ratio and random seed were applied consistently across landmark cohorts to ensure comparability. Probability calibration was performed using isotonic regression fitted exclusively on the calibration set. The test set was held out for final evaluation and was not used for hyperparameter tuning, calibration, or threshold selection.

### Fixed-policy screening evaluation

A fixed screening policy was defined by setting a target specificity of 90% across all models. For each model, the probability cutoff corresponding to this specificity was determined on the calibration set and then applied unchanged to the held-out test set. Because predicted probability scales differ across models, fixing the specificity target rather than a single numerical cutoff ensures that all models operate under an equivalent clinical constraint: the same false positive rate, regardless of how each model scores patients. This approach reflects real-world screening scenarios in which the acceptable rate of false positives is set by clinical or resource considerations, and enables direct assessment of how successive information layers improve DKD detection under a consistent policy.

### Model evaluation and statistical analysis

Discrimination was assessed using the Area Under the Receiver Operating Characteristic Curve (AUROC) and the area under the precision-recall curve (AUPRC), with 95% confidence intervals estimated via nonparametric bootstrapping (n = 2,000 iterations). Pairwise comparisons of discrimination between consecutive models were performed using DeLong tests.^28^

Screening performance at the 90% specificity threshold was summarized using sensitivity (i.e., Recall or True Positive Rate), specificity, positive predictive value (PPV), and negative predictive value (NPV). Individual-level classification transitions between models were quantified, and marginal benefit and harm were assessed by comparing false negative (FN) to true positive (TP) and false positive (FP) to true negative (TN) transitions. Paired comparisons were evaluated using McNemar tests.^29^

The decision curve analysis (DCA) was performed to assess clinical utility across a range of threshold probabilities. The primary comparison evaluated M4 versus M5 among individuals missed by M4, assessing whether the addition of SDOH features provide incremental clinical utility in this subgroup. DCA across the full test population for all model transitions is reported in the Supplement.

### Clinical observability analysis

To quantify clinical observability prior to the 1-year landmark, we defined a domain coverage score as the number of general clinical monitoring domains with at least one recorded measurement. Using measurement count features derived exclusively from data preceding the landmark, eight domains were evaluated: glycemic monitoring (HbA1c, random glucose), blood pressure (SBP, DBP), lipids (LDL-C, HDL-C, triglycerides, total cholesterol), electrolytes (sodium, potassium, chloride, bicarbonate), liver and protein markers (ALT, AST, ALP, albumin, total bilirubin, direct bilirubin, total protein), hematology (hemoglobin), anthropometrics (weight), and vitals (heart rate). Kidney monitoring was excluded to avoid conflating general healthcare engagement with outcome-defining renal biomarkers. Among individuals classified as FN by M4, those reclassified as TP by M5 were designated as rescued, and those remaining FN were designated as not rescued. Participants were grouped into observability deciles, and rescue probability within each decile was estimated with 95% confidence intervals.

### SHAP-based dominance phenotype analyses

We used SHAP (SHapley Additive exPlanations) values to identify contexts in which genetic and social features contributed most strongly to DKD risk prediction. Dominance phenotypes were then defined based on the relative contribution of each feature domain to total model attribution in M5. Features were assigned to thirteen domains: demographics, anthropometrics, vitals, glycemic labs, non-glycemic labs, kidney markers, monitoring patterns, abnormality flags, comorbidity, family history, treatment, social access, and genetics. Genetics comprised all PRS features. Social access comprised SDOH and survey-derived features, including insurance, employment, income, housing, race, marital status, and substance use. Monitoring patterns comprised measurement count and recency features. Abnormality flags comprised ever-abnormal and fraction-abnormal features across biomarker domains.

For each individual in the test set, domain-level attribution was quantified as the fraction of total absolute SHAP values attributable to features within that domain. Genetics-dominant individuals were defined as those in the highest quintile of genetic feature attribution (top 20%). Social-dominant individuals were defined as those in the highest quintile of social access feature attribution (top 20%). The top 20% cutoff was chosen as a simple, prespecified percentile to identify individuals in whom a given domain contributed disproportionately to model predictions.

For each dominance group, mean fractional domain contributions were compared between dominant and non-dominant individuals across all domains. Differences in mean fractional contribution (Δ) were computed as the dominant-group mean minus the non-dominant-group mean, with positive values indicating relatively greater reliance on a domain in dominant individuals. For genetics-dominant individuals, PRS-level contributions were further examined by comparing the mean absolute SHAP value of each PRS between dominant and non-dominant individuals. The ratio of these values was used to quantify relative enrichment.

### External validation

External validation was conducted in the Bio*Me* Biobank using harmonized phenotype definitions, the same one-year landmark definition, and identical feature construction pipelines. Model parameters and screening thresholds learned in the development cohort were applied without retraining to assess strict transportability across healthcare systems with differing population characteristics and care patterns. Discrimination, calibration, screening performance, and marginal benefit analyses were evaluated. As a sensitivity analysis, thresholds were independently relearned in Bio*Me* at 90% specificity to assess model performance under an equivalent screening policy. Because SDOH features were derived from survey instruments administered as part of the AoU Research Program and were not available in Bio*Me*, external validation was restricted to models M1 through M4. Algorithmic sensitivity analyses in Bio*Me* evaluated XGBoost, Random Forest, and Logistic Regression on the M4 feature set.

### Algorithmic sensitivity analyses

To assess robustness to model choice, algorithmic sensitivity analyses were conducted at the one-year landmark using logistic regression (LR) and random forest (RF) classifiers trained on the full feature set corresponding to M5. Both algorithms used the same cohort, outcome definition, and identical 60%, 20%, and 20% training, calibration, and test splits as the primary XGBoost models.

For LR, predictors were standardized following median imputation with missingness indicators, and models were fit using L2 regularization with the regularization strength optimized via Bayesian optimization (Optuna) with five-fold stratified cross-validation. RF models were trained with hyperparameters including the number of trees, maximum depth, minimum samples per leaf, and feature sampling fraction optimized using the same Bayesian optimization framework. Both LR and RF models were calibrated using isotonic regression fitted on the calibration set. For each algorithm, an operating threshold was selected on the calibration set to achieve approximately 90% specificity, matching the fixed screening policy used for the primary analyses, and applied unchanged to the held-out test set. Discrimination, calibration, and screening performance were evaluated using the same metrics as the primary models.

## Results

### Study cohort

A total of 39,431 individuals with established T2D were included from the AoU Research Program after applying the one-year landmark eligibility criteria. Of these, 16,193 (41.1%) developed incident DKD and 23,238 remained DKD-free over the observation period. The one-year landmark defined the prediction time point at which features were assessed; individuals were then followed forward from that point for outcome ascertainment. The median follow-up after the landmark date was 7.7 years (interquartile range [IQR] 4.7-12.3). Baseline demographic and clinical characteristics are summarized in Table 1. Compared with controls, individuals who developed DKD were older at T2D diagnosis, had higher HbA1c and UACR, lower eGFR, and a greater prevalence of hypertension, coronary artery disease, heart failure, and prior acute kidney injury. Exposure to nephroprotective agents, including SGLT2 inhibitors and GLP-1 receptor agonists, was low in both groups, reflecting the early study period.

**Table 1.** Baseline characteristics of the DKD cohorts.

| Characteristic | All of Us |  |  | BioMe |  |  |
| --- | --- | --- | --- | --- | --- | --- |
|  | Controls<br>(N= 23,238) | Cases<br>(N= 16,193) | p-value | Controls<br>(N=6,253) | Cases<br>(N=3,565) | p-value |
| Age at T2D diagnosis, years | 51.78 ± 13.73 | 55.85 ± 12.28 | <0.001 | 53.47 ± 13.72 | 56.23 ± 11.71 | <0.001 |
| HbA1c, % | 6.92 ± 1.78 | 7.18 ± 1.80 | <0.001 | 6.78 ± 1.48 | 7.44 ± 1.89 | <0.001 |
| eGFR, mL/min/1.73 m<sup>2</sup> | 94.22 ± 16.89 | 81.55 ± 19.63 | <0.001 | 95.20 ± 18.00 | 89.99 ± 18.72 | <0.001 |
| Creatinine, mg/dL | 0.79 ± 0.29 | 0.95 ± 0.59 | <0.001 | 0.82 ± 0.21 | 0.87 ± 0.33 | <0.001 |
| UACR, mg/g | 7.10 (4.31–14.04) | 13.86 (6.03–40.04) | <0.001 | 7.00 (4.00–14.00) | 9.00 (5.00–16.00) | <0.001 |
| Urine albumin | 0.99 (0.50–1.80) | 1.55 (0.70–5.60) | <0.001 | 1.00 (0.47–3.20) | 1.06 (0.52–3.02) | 0.161 |
| SBP, mmHg | 127.49 ± 13.64 | 130.28 ± 14.57 | <0.001 | 127.74 ± 17.37 | 130.40 ± 18.29 | <0.001 |
| DBP, mmHg | 76.69 ± 8.96 | 76.74 ± 9.01 | 0.661 | 75.90 ± 10.49 | 75.78 ± 10.62 | 0.439 |
| LDL, mg/dL | 104.87 ± 34.23 | 105.30 ± 35.18 | 0.664 | 99.24 ± 36.98 | 97.06 ± 37.82 | 0.004 |
| Triglycerides, mg/dL | 129.00 (89.00–185.00) | 141.67 (99.00–206.00) | <0.001 | 123.00 (85.00–179.00) | 134.00 (93.00–196.00) | <0.001 |
| Male sex, % | 39.3% | 45.3% | <0.001 | 43.3% | 39.6% | <0.001 |
| Hypertension, % | 58.6% | 69.8% | <0.001 | 37.4% | 33.6% | <0.001 |
| CAD, % | 12.0% | 16.9% | <0.001 | 8.1% | 5.7% | <0.001 |
| Heart failure, % | 6.7% | 11.0% | <0.001 | 2.5% | 2.1% | 0.225 |
| Atrial fibrillation, % | 5.3% | 7.2% | <0.001 | 2.4% | 1.7% | 0.022 |
| Obesity, % | 40.0% | 38.2% | <0.001 | 21.1% | 12.5% | <0.001 |
| AKI history, % | 3.7% | 8.6% | <0.001 | 0.3% | 0.4% | 0.326 |
| ACE inhibitors exposure, % | 23.3% | 31.7% | <0.001 | 17.6% | 18.2% | 0.503 |
| ARB exposure, % | 10.8% | 13.6% | <0.001 | 10.3% | 7.8% | <0.001 |
| SGLT2 inhibitors exposure, % | 1.2% | 1.1% | 0.322 | 1.4% | 0.8% | 0.013 |
| GLP-1RA exposure, % | 2.5% | 2.1% | 0.018 | 3.8% | 1.6% | <0.001 |
| Insulin exposure, % | 8.0% | 10.0% | <0.001 | 14.5% | 11.6% | <0.001 |
| Statin exposure, % | 33.1% | 40.9% | <0.001 | 30.4% | 24.6% | <0.001 |

† Values are mean ± SD or median (IQR) unless otherwise indicated.
T2D, type 2 diabetes; HbA1c, hemoglobin A1c; eGFR, estimated glomerular filtration rate; UACR, urine albumin-to-creatinine ratio; SBP, systolic blood pressure; DBP, diastolic blood pressure; LDL, low-density lipoprotein cholesterol; CAD, coronary artery disease; AKI, acute kidney injury; ACE, angiotensin-converting enzyme; ARB, angiotensin receptor blocker; SGLT2, sodium-glucose cotransporter 2; GLP-1RA, glucagon-like peptide-1 receptor agonist.

External validation was conducted in the Bio*Me* Biobank, which included 9,818 individuals with T2D who met the same one-year landmark eligibility criteria, of whom 3,565 (36.3%) developed incident DKD during follow-up. Relative to AoU, Bio*Me* showed lower baseline burden across several domains, including lower prevalences of hypertension, obesity, coronary artery disease, and prior acute kidney injury, along with more favorable kidney and lipid measures. Notably, Bio*Me* cases had higher mean eGFR and lower median UACR than AoU cases. These differences may reflect population, healthcare system, and data-capture differences between cohorts. Baseline characteristics are summarized in Table 1.

### Model performance across information layers

Model discrimination improved consistently with the addition of successive information layers (Supplementary Table S4). M1, incorporating intrinsic risk features alone, achieved an AUROC of 0.673 (95% CI: 0.661–0.684) and an AUPRC of 0.565 (95% CI: 0.549–0.582). The addition of laboratory snapshot features in M2 produced the largest single gain, with AUROC increasing to 0.763 (95% CI: 0.753–0.774) and AUPRC to 0.693 (95% CI: 0.677–0.708; ΔAUROC 0.090, DeLong p <1×10□^3^□□).

Subsequent layers yielded additional but progressively smaller gains. M3 achieved an AUROC of 0.764 (95% CI: 0.754–0.774) and AUPRC of 0.696 (95% CI: 0.681–0.711), with a statistically significant improvement in classification under the fixed screening policy (McNemar p = 0.0005) but no significant change in overall discrimination (ΔAUROC 0.001, DeLong p = 0.50). M4 demonstrated a more substantial improvement in discrimination, with AUROC of 0.786 (95% CI: 0.775–0.795) and AUPRC of 0.726 (95% CI: 0.711–0.741; ΔAUROC 0.021, DeLong p = 3.9×10□^12^). M5 achieved the highest overall discrimination, with AUROC of 0.797 (95% CI: 0.787–0.807) and AUPRC of 0.737 (95% CI: 0.722–0.752; ΔAUROC 0.011, DeLong p = 5.4×10□□). Calibration was good across all models following isotonic regression, with predicted probabilities closely tracking observed event rates (Figure 1c).

**Figure 1.**
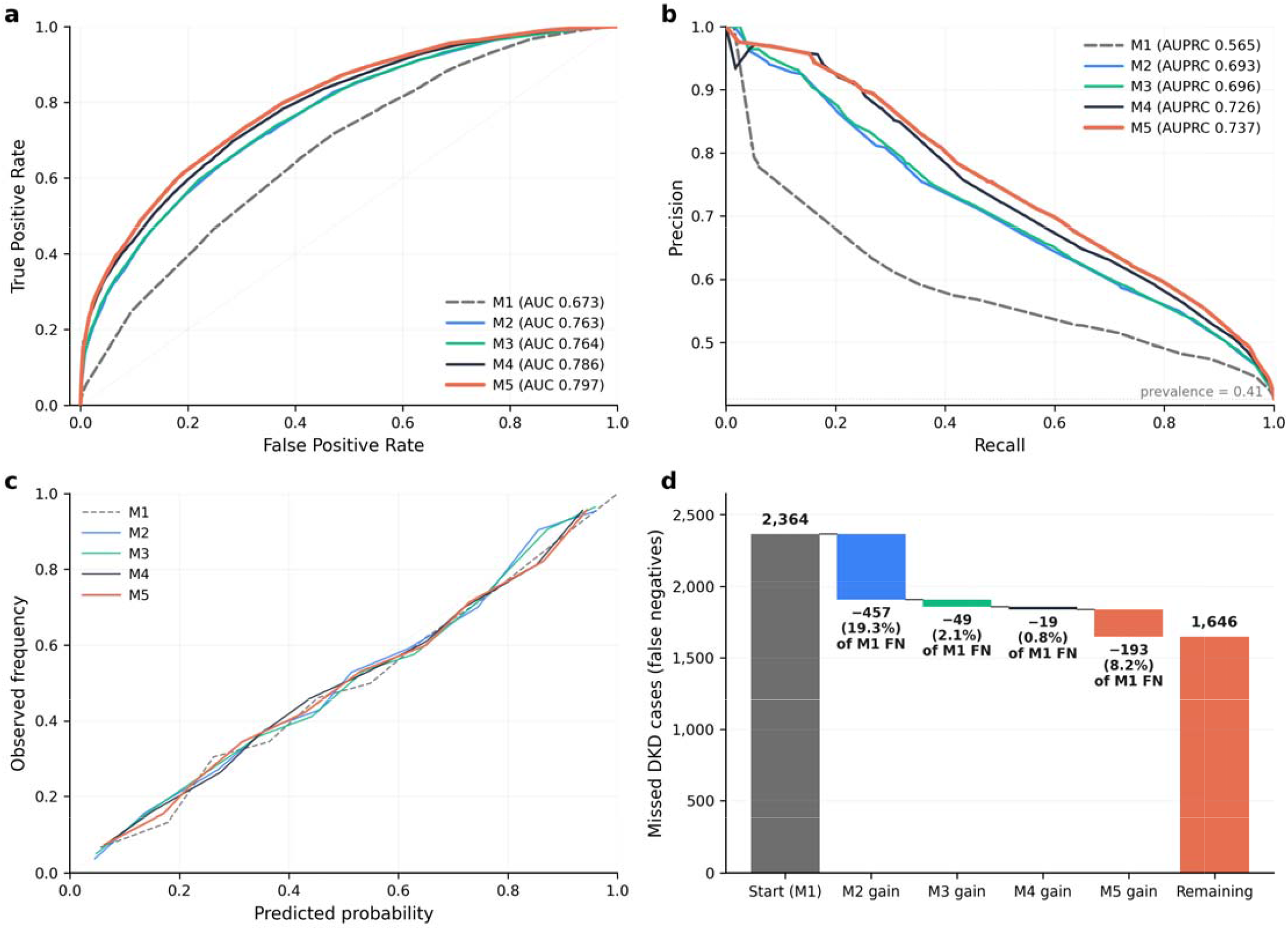
Discrimination, calibration, and cumulative recovery of missed DKD cases at the 1-year landmark. **(a)** Receiver operating characteristic (ROC) curves for Models M1–M5, demonstrating progressive improvement in discrimination with the sequential addition of information layers. **(b)** Precision–recall (PR) curves for Models M1–M5, with the horizontal dashed line indicating outcome prevalence at the 1-year landmark (41.1%). **(c)** Calibration plots showing observed versus predicted risk after isotonic regression calibration, indicating good agreement across models. **(d)** Waterfall plot illustrating cumulative recovery of missed DKD cases (false negatives) under model-specific screening thresholds selected to achieve approximately 90% specificity on the calibration set. Bars represent the incremental number and percentage of cases rescued by each successive model layer, expressed as a proportion of false negatives under M1. Layered modeling recovered 30.4% of cases missed by the base intrinsic risk model. M1, intrinsic risk; M2, M1 plus laboratory snapshots; M3, M2 plus medication exposure; M4, M3 plus longitudinal care trajectories; M5, M4 plus social determinants of health.

### Fixed-policy screening performance and calibration

Under the fixed screening policy, sensitivity increased from 0.27 (95% CI 0.26–0.28) in M1 to 0.49 (95% CI 0.47–0.51) in M5, representing a near-doubling of case detection at equivalent specificity, with PPV improving from 0.63 (95% CI 0.61–0.66) to 0.75 (95% CI 0.73–0.77). Full screening performance metrics with confidence intervals are reported in Table 2.

**Table 2.** Screening performance of models M1–M5 under a fixed clinical screening policy (1-year landmark)

| Model | Sensitivity<br>(95% CI) | Specificity<br>(95% CI) | PPV<br>(95% CI) | NPV<br>(95% CI) | TP | FP | TN | FN |
| --- | --- | --- | --- | --- | --- | --- | --- | --- |
| M1 | 0.27 (0.26–0.28) | 0.89 (0.88–0.90) | 0.63 (0.61–0.66) | 0.64 (0.62–0.65) | 875 | 512 | 4,136 | 2,364 |
| M2 | 0.41 (0.39–0.43) | 0.90 (0.89–0.90) | 0.73 (0.71–0.75) | 0.69 (0.67–0.70) | 1,332 | 488 | 4,160 | 1,907 |
| M3 | 0.43 (0.41–0.44) | 0.89 (0.88–0.90) | 0.73 (0.71–0.75) | 0.69 (0.68–0.70) | 1,381 | 518 | 4,130 | 1,858 |
| M4 | 0.43 (0.42–0.45) | 0.90 (0.89–0.91) | 0.76 (0.74–0.78) | 0.70 (0.68–0.71) | 1,400 | 448 | 4,200 | 1,839 |
| M5 | 0.49 (0.47–0.51) | 0.89 (0.88–0.90) | 0.75 (0.73–0.77) | 0.71 (0.70–0.73) | 1,593 | 530 | 4,118 | 1,646 |

† A probability threshold corresponding to 90% specificity was selected independently for each model on the calibration set and applied unchanged to the held-out test set.
‡ All models were evaluated on identical test individuals (n = 7,887), enabling paired comparison across models.
§ Confidence intervals were estimated using nonparametric bootstrapping (n = 2,000 iterations).
¶ M1, intrinsic risk features; M2, laboratory snapshot features added; M3, medication exposure features added; M4, longitudinal trajectory features added; M5, social determinants of health added.
PPV, positive predictive value; NPV, negative predictive value; TP, true positive; FP, false positive; TN, true negative; FN, false negative.

The largest single gain occurred at the M1→M2 transition, where laboratory snapshot features recovered 740 previously missed DKD cases, representing a 19.4% reduction in false negatives relative to M1 (McNemar p = 3.2×10□^2^□). Subsequent layers contributed additional but progressively smaller gains: medication features (M2→M3) reduced remaining false negatives by 2.6% (net 49; McNemar p = 0.0005), longitudinal trajectory features (M3→M4) by 1.0% (net 19; McNemar p = 0.084), and SDOH (M4→M5) by 10.1% (net 193; McNemar p = 1.6×10□^2^□). Across all transitions, gains were driven predominantly by FN→TP conversions with stable false positive burden. Cumulatively, layered modeling reduced missed DKD cases by 30.4% relative to M1 under the fixed screening policy (Figure 1d). Complete transition counts and paired statistical comparisons are provided in Supplementary Table S4.

### Characteristics of individuals rescued by successive information layers

To characterize which individuals benefited from successive information layers, we compared individuals transitioning from FN to TP at each model transition with those remaining FN.

Medication exposure and treatment intensity (M2→M3). Individuals rescued by the addition of medication features had greater cumulative exposure to glucose-lowering and cardiovascular therapies, including sulfonylureas, metformin, statins, ACE inhibitors, beta blockers, and insulin, consistent with longer disease duration and higher cardiometabolic burden. These patterns are summarized in Figure 2a.

**Figure 2.**
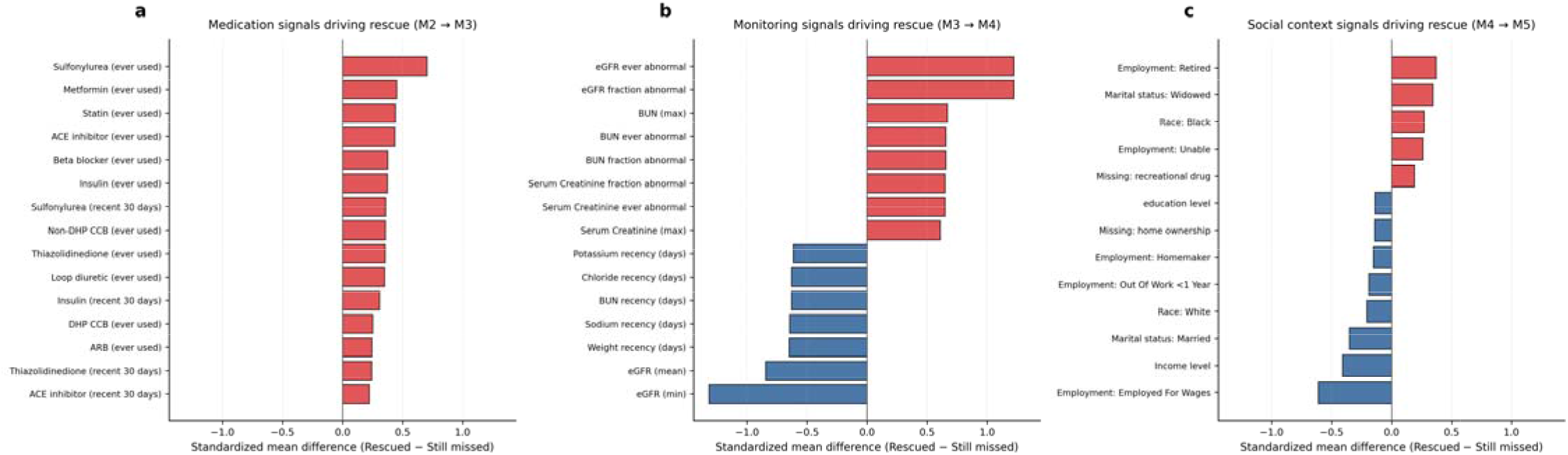
Characteristics of individuals rescued by successive information layers at the 1-year landmark. Standardized mean differences comparing individuals transitioning from false negative to true positive at each model layer with those remaining false negative. Positive values indicate enrichment among rescued individuals; negative values indicate enrichment among individuals remaining missed. **(a)** Medication exposure patterns distinguishing cases newly detected by M3 among individuals missed by M2, reflecting higher treatment intensity and cardiometabolic disease burden among rescued individuals. **(b)** Care-aware longitudinal features distinguishing cases newly detected by M4 among individuals missed by M3, reflecting greater biological instability and denser clinical monitoring among rescued individuals. **(c)** Social and access-related characteristics distinguishing cases newly detected by M5 among individuals missed by M4, reflecting differences in insurance status, employment, and social engagement. ACE, angiotensin-converting enzyme; ARB, angiotensin receptor blocker; BUN, blood urea nitrogen; CCB, calcium channel blocker; DHP, dihydropyridine; eGFR, estimated glomerular filtration rate; FPG, fasting plasma glucose; GLP-1RA, glucagon-like peptide-1 receptor agonist; SGLT2, sodium–glucose cotransporter 2; UACR, urine albumin-to-creatinine ratio.

Longitudinal trajectory signals and clinical monitoring (M3→M4). Individuals rescued by longitudinal trajectory features showed two complementary patterns. First, they exhibited greater biological instability in kidney markers, reflected by higher proportions of ever-abnormal and repeatedly abnormal eGFR and BUN measurements over time, indicating persistent rather than transient kidney dysfunction. Second, individuals remaining FN were distinguished by longer recency intervals across electrolyte, renal, and anthropometric monitoring domains, indicating sparser longitudinal records prior to the landmark. Rescue at this transition, therefore, required both the presence of biological instability and sufficient data density to render that instability observable within the clinical record (Figure 2b).

Social and access-related features (M4→M5). Individuals rescued by the addition of SDOH represented a distinct subgroup characterized by differences in healthcare access and engagement. Rescued individuals were more likely to report retirement or inability to work, were more commonly identified as Black race, and had lower income levels relative to those remaining missed. Individuals remaining FN were more likely to be employed and had higher income levels. These patterns are summarized in Figure 2c.

### Clinical observability and SDOH rescue

Among individuals classified as FN by M4, rescue probability by M5 declined monotonically with increasing clinical observability. Rescue probability was highest in the lowest observability decile at 29.7% (95% CI: 24.5%–35.4%), decreased across intermediate deciles to approximately 10%–20%, and reached its lowest point in the highest decile at 9.6% (26/270; 95% CI: 6.7%–13.7%). DCA restricted to individuals who remained FN under M4 demonstrated higher net benefit for M5 across clinically relevant threshold probabilities, supporting the targeted clinical utility of SDOH augmentation in this low-observability subgroup (Figure 3a, 3c).

**Figure 3.**
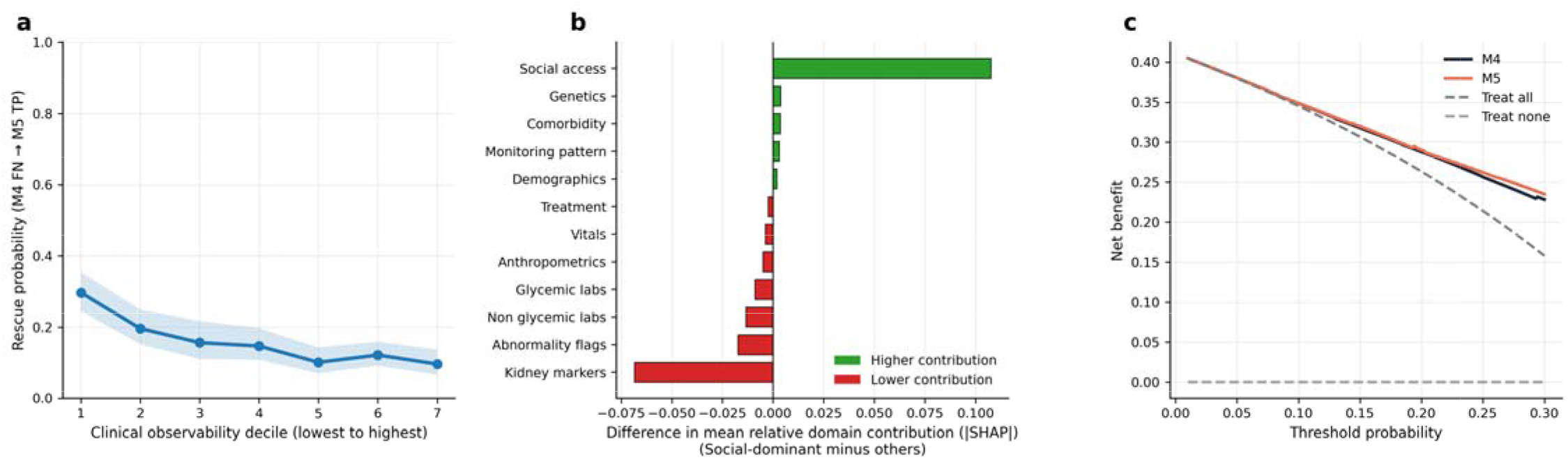
Clinical observability, social dominance, and decision utility at the 1-year landmark. **(a)** Rescue probability by M5 as a function of clinical observability decile among individuals missed by M4. Observability was quantified as the number of general clinical monitoring domains with at least one recorded measurement prior to the landmark, excluding kidney-specific domains. Rescue probability declined monotonically from 29.7% (95% CI 24.5–35.4%) in the lowest decile to 9.6% (95% CI 6.7–13.7%) in the highest, indicating that SDOH features preferentially recovered individuals with sparse clinical records. **(b)** Difference in mean fractional absolute SHAP contribution across feature domains comparing social-dominant individuals (top 20% by relative social access contribution in M5) with all others. Positive values indicate domains contributing more strongly among social-dominant individuals. Social access showed the largest relative enrichment, while kidney markers and abnormality flags contributed less, indicating that contextual features compensate when the physiologic monitoring signal is limited. **(c)** Decision curve analysis comparing M4 and M5 among individuals missed by M4, demonstrating higher net benefit for M5 across clinically relevant threshold probabilities. DKD, diabetic kidney disease; FN, false negative; SDOH, social determinants of health; SHAP, SHapley Additive exPlanations; TP, true positive.

### Social dominance phenotype and model attribution

Among the 7,887 individuals in the test set, 1,578 (20.0%) were classified as social-dominant based on social access feature attribution in M5. Relative to all others, social-dominant individuals exhibited a systematic redistribution of predictive signal away from downstream clinical domains toward upstream and less care-dependent domains. Kidney markers showed the largest relative decrease in mean fractional attribution (12.8% vs 19.7%; Δ = -0.068, 95% CI -0.071 to -0.065), followed by lab abnormality flags (Δ = -0.017, 95% CI -0.019 to -0.016) and non-glycemic laboratory features (Δ = -0.013, 95% CI -0.015 to -0.012). Social access showed the largest relative enrichment (24.2% vs 13.4%; Δ = +0.107, 95% CI +0.105 to +0.110), indicating that contextual features compensated for the limited physiologic signal in this subgroup (Figure 3b).

### Genetic dominance phenotype and polygenic drivers

Among the 7,887 individuals in the test set, 1,578 (20.0%) were classified as genetics-dominant based on genetic feature attribution in M5. Relative to all others, genetics-dominant individuals exhibited a similar pattern of redistribution away from downstream clinical domains. Kidney markers showed the largest relative decrease in mean fractional attribution (14.0% vs. 19.3%, Δ=−0.053, 95% CI -0.056 to -0.050), followed by lab abnormality flags (Δ=−0.014, 95% CI -0.016 to - 0.013). Genetic features showed the largest relative enrichment (9.5% vs. 4.3%, Δ= +0.052, 95% CI +0.051 to +0.053), with social access also contributing more in this subgroup (Δ=+0.014, 95% CI +0.011 to +0.017). These patterns suggest that genetic and contextual signals contributed more when downstream clinical information was less prominent (Figure 4a).

**Figure 4.**
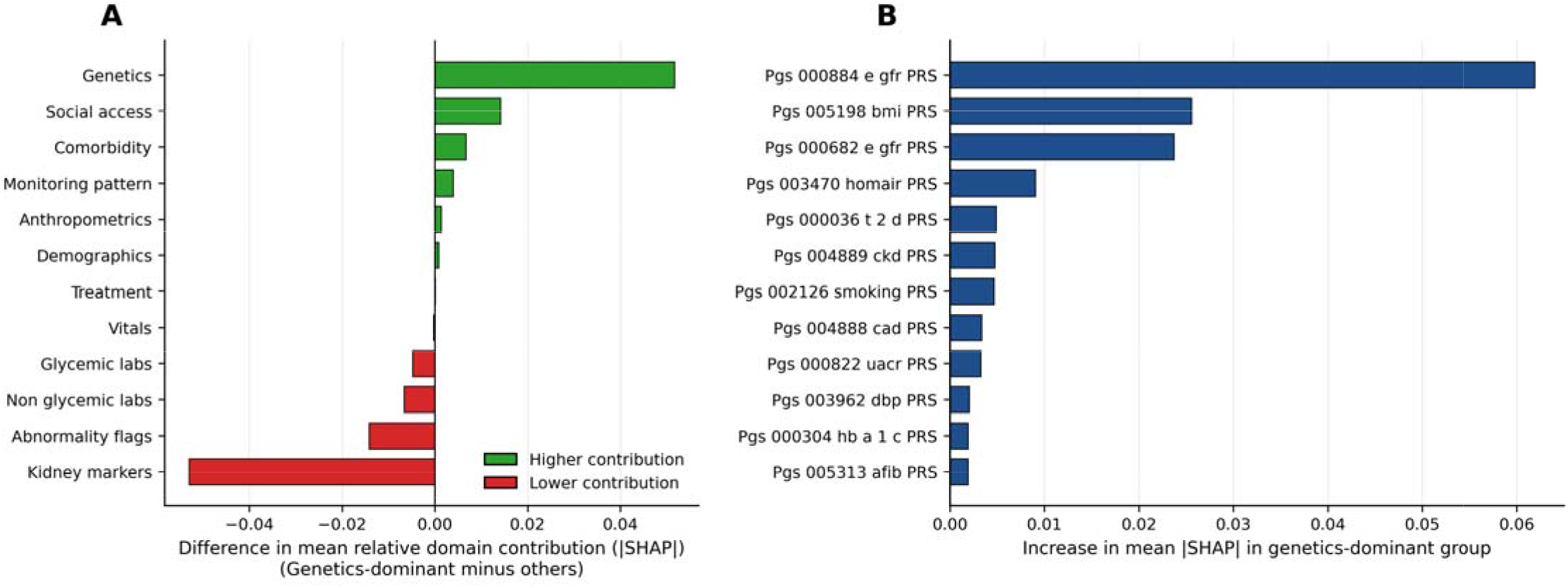
Genetic dominance phenotype and polygenic drivers of model contribution at the 1-year landmark. **(a)** Difference in mean fractional absolute SHAP contribution across feature domains comparing genetics-dominant individuals (top 20% by relative genetic contribution in M5) with all others. Positive values indicate domains contributing more strongly among genetics-dominant individuals. Genetic features showed the largest relative enrichment, while care intensity and kidney markers contributed less, indicating that genetic information is preferentially leveraged when the downstream clinical signal is sparse. **(b)** Individual polygenic risk scores ranked by increase in mean absolute SHAP value among genetics-dominant individuals relative to others. Renal function and metabolic PRS — including eGFR (PGS000884, PGS000682) and BMI (PGS005198) — showed the largest incremental contribution, identifying specific genetic pathways that drive risk stratification in this subgroup. BMI, body mass index; eGFR, estimated glomerular filtration rate; HOMA-B, homeostatic model assessment of beta-cell function; HOMA-IR, homeostatic model assessment of insulin resistance; PRS, polygenic risk score; SHAP, SHapley Additive exPlanations; T2D, type 2 diabetes; UACR, urine albumin-to-creatinine ratio.

At the feature level, several renal and metabolic PRS contributed more strongly in genetics-dominant individuals than in all others. The eGFR PRS (PGS000884) showed the largest increase in mean absolute SHAP value (Δ = +0.062, 95% CI +0.059 to +0.065) and was enriched 3.0-fold relative to the non-dominant group. The BMI PRS (PGS005198) was similarly enriched (3.1-fold; Δ = +0.026, 95% CI +0.023 to +0.028), followed by a second eGFR PRS (PGS000682; 2.5-fold; Δ = +0.024, 95% CI +0.021 to +0.026). PRS for insulin resistance (HOMA-IR; 1.9-fold; Δ = +0.009, 95% CI +0.008 to +0.010), T2D (1.5-fold; Δ = +0.005, 95% CI +0.004 to +0.006), and CKD (1.7-fold; Δ = +0.0048, 95% CI +0.0042 to +0.0053) also showed stronger contributions in this subgroup. These findings highlight specific renal and metabolic PRS that were most strongly emphasized in individuals whose predictions were genetics-driven (Figure 4b).

### Algorithmic sensitivity analysis

To assess robustness to model choice, LR and RF were evaluated on the same held-out test set as the primary XGB models, using the full M5 feature set and thresholds selected on the calibration set to achieve approximately 90% specificity. At this fixed specificity, XGB achieved the highest sensitivity at 0.49 (95% CI: 0.47–0.51), compared with 0.39 (95% CI: 0.38–0.41) for both LR and RF. Specificity was broadly comparable across algorithms on the same test individuals, ranging from 0.89 (95% CI: 0.88–0.90) for XGB to 0.93 (95% CI: 0.93–0.94) for LR. PPV followed an inverse pattern, with LR achieving 0.80 (95% CI: 0.78–0.82) and RF 0.77 (95% CI: 0.75–0.79) compared with 0.75 (95% CI: 0.73–0.77) for XGB, reflecting the higher specificity operating points of the linear and ensemble models.

Discrimination also differed across algorithms on the held-out test set, with XGB achieving the highest AUROC at 0.797 (95% CI: 0.787–0.807), followed by LR at 0.788 (95% CI: 0.778–0.798) and RF at 0.780 (95% CI: 0.770–0.791). Paired DeLong tests confirmed statistically significant differences between XGB and both LR (ΔAUROC=0.009, p=0.0006) and RF (ΔAUROC=0.016, p=2.6×10□^13^). Complete performance metrics are provided in Supplementary Table S5.

### External validation

Model parameters and screening thresholds learned in the AoU development cohort were applied without retraining to the Bio*Me* cohort at the one-year landmark. External validation was restricted to M1 through M4, as SDOH survey features were not available in Bio*Me*. Detailed model specifications, including optimized hyperparameters, are provided in Supplementary Table S6.

Discrimination was attenuated relative to AoU across all model layers, with XGB M4 achieving an AUROC of 0.659 (95% CI: 0.648–0.670) compared with 0.781 in AoU. The relative ordering of information layers was fully preserved: M1 achieved an AUROC of 0.546 (95% CI: 0.534–0.558), with the largest single gain at M1→M2 (ΔAUROC 0.075, DeLong p < 1×10□^33^), followed by M3→M4 (ΔAUROC 0.047, DeLong p < 1×10□^2^□). The M2→M3 transition was statistically significant but modest (ΔAUROC 0.008, DeLong p = 0.003), consistent with AoU.

Application of AoU-derived thresholds to Bio*Me* revealed a systematic upward shift in the probability cutoffs required to achieve 90% specificity, most pronounced for XGB M4 (0.812 vs. 0.571 in AoU) and LR (0.892 vs. 0.595), and smaller for RF (0.624 vs. 0.575). Under Bio*Me*-native 90% specificity thresholds, XGB M4 achieved a sensitivity of 0.171 (PPV 0.529), RF achieved a sensitivity of 0.153 (PPV 0.535), and LR achieved a sensitivity of 0.096 (PPV 0.568). Across all three algorithms, discrimination ordering was consistent with AoU: XGB M4 achieved the highest AUROC (0.659), followed by RF (0.646) and LR (0.610), with all pairwise differences statistically significant (DeLong p < 5×10□□ for all comparisons). Performance is summarized in Supplementary Table S7.

The attenuation in Bio*Me* should be interpreted in the context of baseline differences between cohorts. As shown in Table 1, Bio*Me* differed from AoU across multiple clinical domains, which may have contributed to lower external performance.

## Discussion

Existing approaches to DKD risk prediction in T2D typically optimize models at a single cross-sectional snapshot and evaluate performance using model-specific thresholds, making it difficult to understand how additional data sources alter real-world screening behavior or which patients benefit when models are expanded. We addressed these limitations by combining two methodological choices that are rarely applied together: a landmark prediction framework that anchors risk assessment to a defined point in the T2D disease course, and a fixed screening policy that holds the false positive rate constant across all model layers. This design directly answers a question that global discrimination metrics cannot address: as more information becomes available about a patient, who gets detected that was previously missed, and why? The answer, across all model transitions, was consistent. Successive information layers recovered distinct subgroups of at-risk individuals rather than redistributing the same cases, and the characteristics of rescued individuals reflected the specific type of information added. This pattern reframes DKD risk detection not as a problem of discrimination alone, but as a problem of risk observability: whether the biological and contextual signals needed to identify an at-risk individual are present and measurable within the clinical record at the time of screening.

The addition of laboratory snapshot features yielded the largest single gain in DKD detection, with sensitivity increasing from 0.27 to 0.41 under the fixed screening policy, driven by net recovery of 457 previously missed cases. This finding is consistent with the established role of glycemic and renal biomarkers in DKD risk assessment and confirms that cross-sectional laboratory data capture the majority of biologically identifiable risk at the 1-year landmark. However, nearly three in five incident DKD cases remained missed after laboratory features were added, underscoring that snapshot-based biological information alone is insufficient for comprehensive screening. The remaining undetected risk was not uniformly distributed but concentrated in individuals whose clinical trajectories, treatment histories, and social contexts differed systematically from those identified by biology alone, motivating the successive information layers.

Incorporation of medication exposure features recovered an additional 130 previously missed cases under the fixed screening policy, with a modest but statistically significant improvement in classification. In this predictive context, medication exposure patterns served as indicators of underlying disease burden and clinical management history. Individuals rescued by this layer were more likely to have cumulative exposure to glucose-lowering and cardiovascular therapies, consistent with longer disease duration and higher cardiometabolic burden. These findings highlight that medication exposure captures complementary information about clinical trajectory not fully reflected in laboratory values alone.

Incorporation of longitudinal trajectory features yielded the second largest gain in DKD detection, recovering 12.5% of cases remaining missed after medication features were added. This improvement reflects a fundamental distinction between what a single laboratory measurement captures and what a series of measurements over time reveals. Rescued individuals exhibited greater biological instability in kidney markers, characterized by higher proportions of persistently and repeatedly abnormal eGFR and BUN measurements, indicating that trajectory-based features encode not just current biological state but the evolution and consistency of dysfunction over time. Critically, rescue at this transition required not only the presence of biological instability but sufficient data density to render that instability observable within the clinical record. Individuals who remained missed after trajectory features were added consistently exhibited sparse longitudinal records across multiple monitoring domains, indicating that risk detectability in EHR-based models is shaped not only by underlying biology but by the extent to which that biology is observed through healthcare interactions. This finding has a direct clinical implication: sparse data in the record does not indicate low risk. It indicates low observability.

Inclusion of SDOH features recovered 19.1% of cases remaining missed after longitudinal trajectory features were added, representing a clinically meaningful gain concentrated in a distinct subgroup rather than broadly distributed across the missed population. Rescued individuals were characterized by lower clinical observability, with rescue probability highest among those with the fewest recorded monitoring domains prior to the landmark and declining monotonically as data density increased. This pattern indicates that SDOH features provided the most incremental value precisely when biological and care trajectory data were sparse, effectively compensating for limited physiologic signal through contextual information about healthcare access and social circumstance. Differences in insurance status, employment, and social isolation among rescued individuals suggest that these were not simply high-risk patients who happen to have social disadvantage, but patients whose risk was not otherwise visible in the clinical record because their engagement with the healthcare system was limited. These findings support a targeted rather than universal role for SDOH in DKD screening, with the greatest utility in settings where traditional clinical data are sparse or unavailable.

Although genetic features contributed modestly to overall discrimination at the population level, they played a disproportionate role in specific clinical contexts. Genetics-dominant individuals, defined as those in the highest quintile of genetic feature attribution, exhibited a systematic redistribution of predictive signal away from downstream care-dependent domains, including kidney markers and lab abnormality flags, toward upstream features, including genetic risk, family history, and SDOH. This pattern mirrors what was observed for social-dominant individuals and points to a shared underlying mechanism: when accumulated clinical data is limited, the model relies more heavily on features that are present regardless of healthcare engagement. At the feature level, renal function and metabolic PRS, particularly eGFR and BMI PRS, showed the largest conditional enrichment in genetics-dominant individuals, suggesting that specific biological pathways captured by these scores are informative precisely in contexts where downstream physiologic manifestations are not yet observable in the clinical record. Together, these findings support a contextual rather than universal role for genetic information in EHR-based DKD screening, complementing rather than replacing longitudinal clinical signals.

To assess whether observed gains were specific to the gradient-boosted tree implementation, we evaluated LR and RF on the full feature set under the same fixed screening policy. While XGB achieved the highest sensitivity and discrimination, both alternative algorithms demonstrated meaningful performance at the same fixed specificity, with consistent ordering across metrics. Notably, LR achieved higher PPV and specificity than XGB at its operating point, reflecting a tradeoff between sensitivity and precision that will depend on the clinical context and the relative costs of missed cases versus false alarms. Across all three algorithms, the ordering of performance was consistent and differences in discrimination were statistically significant, indicating that observed gains are driven primarily by feature content and information layering rather than algorithm-specific properties. This supports the generalizability of the proposed framework across modeling approaches and suggests that the layered feature structure, rather than any particular implementation, is the primary driver of screening performance.

The generalizability of this framework was assessed through external validation in the Bio*Me* Biobank, an independent EHR-linked biobank with a diverse demographic composition reflective of a major urban healthcare system. Discrimination was attenuated in Bio*Me* relative to the development cohort, consistent with known challenges of EHR-based model transportability across healthcare systems with differing care patterns, coding practices, and data density. Systematic differences in comorbidity and medication coding between the two systems, reflected in markedly lower prevalences of hypertension, obesity, and AKI history in Bio*Me*, suggest that both population differences and sparser clinical recording may have contributed to attenuated performance in the external cohort. This is consistent with the observability framing central to this study: if risk-relevant clinical information is less completely recorded, models trained on richer data will underperform not because the biology differs, but because the record is less complete. Importantly, the relative ordering of information layers was fully preserved across both cohorts, and the dominant gains at M1 to M2 and M3 to M4 remained statistically significant in Bio*Me*, indicating that the layered framework generalizes in structure even when absolute discrimination is attenuated. A systematic upward shift in predicted probability distributions was observed in Bio*Me*, requiring substantially higher thresholds to achieve equivalent specificity, which underscores the need for recalibration before deployment in a new healthcare system. The medication layer contributed minimally to discrimination in Bio*Me*, suggesting that treatment-based signals may be more susceptible to cross-system coding differences than biological or trajectory-based features. Together, these findings support the transportability of the layered framework as an analytical structure while highlighting that data completeness assessment and site-specific calibration are prerequisites for clinical application.

Some limitations warrant consideration. First, DKD was defined as a composite phenotype based on persistent laboratory abnormalities and diagnosis codes, without subclassification by severity or progression stage. This reflects both the constraints of EHR data and the study’s focus on screening rather than prognostication, but may result in heterogeneity within the outcome that limits interpretation of which disease trajectories are being detected. Second, the landmark framework requires a minimum of one year of follow-up after T2D diagnosis, which excludes early progressors and limits applicability to individuals with rapidly developing kidney involvement. Third, SDOH features were derived from survey instruments administered as part of the AoU research program and may not be routinely available in standard clinical settings, limiting the immediate deployability of the full M5 model in real-world screening pipelines. Fourth, although the AoU Research Program is designed to oversample underrepresented populations, patterns of healthcare engagement and data availability in this cohort may differ from other healthcare systems, and the generalizability of observability-related findings to settings with different monitoring practices warrants further investigation. Fifth, external validation in Bio*Me* was limited to M1 through M4 because SDOH survey features were not available. Moreover, the lower external performance observed in Bio*Me* may reflect several differences between cohorts, including event rate, baseline clinical characteristics, predictor distributions, and EHR data capture, rather than any single factor alone. These sources of heterogeneity could not be formally disentangled in the present study. t

This study demonstrates that DKD risk detection in T2D can be substantially improved by integrating complementary information layers under a fixed clinical screening policy, with a cumulative recovery of 30.4% of cases missed by the base intrinsic risk model. Gains arose from distinct domains that identified at-risk individuals in different clinical contexts: biologically unstable patients with dense longitudinal monitoring, individuals with high treatment intensity reflecting advanced cardiometabolic burden, and those with limited clinical observability whose risk was rendered visible through contextual and genetic information. The layered landmark framework introduced here offers a principled approach to EHR-based DKD screening that aligns model evaluation with clinical deployment realities, accounts for the role of data availability in risk detection, and provides a foundation for adaptive screening strategies that can be tailored to the information available for each patient at the time of risk assessment. External validation in the Bio*Me* Biobank confirmed that the relative ordering of information layers generalizes across healthcare systems, while the systematic attenuation of discrimination and shift in predicted probability distributions underscore the importance of recalibration and data completeness assessment before clinical deployment.

## Acknowledgments

We gratefully acknowledge All of Us participants for their contributions, without whom this research would not have been possible. We also thank the National Institutes of Health’s All of Us Research Program for making available the participant data examined in this study.

The Mount Sinai Bio*Me* Biobank is supported by The Andrea and Charles Bronfman Philanthropies and by Federal funds from the NIH (U01HG00638001; U01HG007417; X01HL134588). We thank all participants and all our recruiters who have assisted and continue to assist in data collection and management. Furthermore, analyses were in part supported through the computational and data resources and staff expertise provided by Scientific Computing and Data at the Icahn School of Medicine at Mount Sinai and by the Clinical and Translational Science Awards (CTSA) grant UL1TR004419 from the National Center for Advancing Translational Sciences. Research reported in this publication was also supported by the Office of Research Infrastructure of the National Institutes of Health under award number S10OD026880 and S10OD030463. The content is solely the responsibility of the authors and does not necessarily represent the official views of the National Institutes of Health.

## Author contributions

A.K. conceived the study, designed the analytical framework, performed the analyses, and drafted the manuscript. Z.W. conducted the Bio*Me* data curation and external validation analyses, contributed to methodological development, and reviewed and revised the manuscript. V.S. contributed to methodological discussions and data curation strategy. H.K.T. contributed to methodological guidance, interpretation of results, and critical review and revision of the manuscript. R.L. contributed to the design and oversight of the Bio*Me* component and to manuscript review and revision. N.L. contributed to study oversight, interpretation of findings, and manuscript review and revision. M.R.I. supervised the study, contributed to study design, methodological guidance, and interpretation of findings, and critically revised the manuscript. All authors reviewed and approved the final manuscript.

## Data availability

The All of Us Research Program data are available to authorized researchers through the All of Us Researcher Workbench, subject to program policies and data access requirements. Bio*Me* data are not publicly available and are accessible only through approved institutional processes and data use agreements.

## Supplementary information

Supplementary information is available for this paper.

