## Supplement for "Where risk becomes visible: a layered fixed-policy framework for diabetic kidney disease screening in type 2 diabetes"

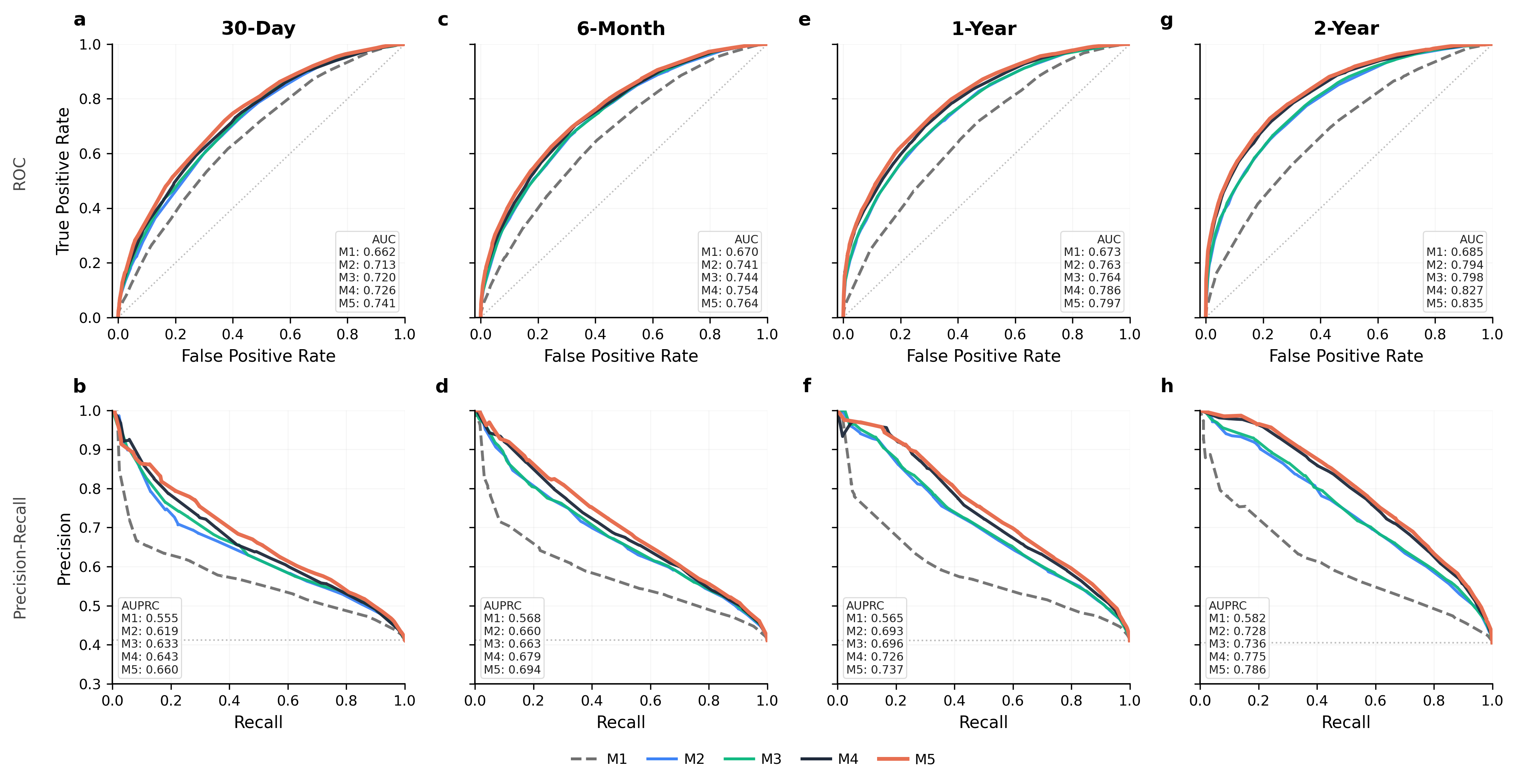

**Supplementary Figure S1. Model discrimination across prediction landmarks.**

ROC (top) and precision–recall (bottom) curves for models M1–M5 evaluated at the 30-day (a, b), 6-month (c, d), 1-year (e, f), and 2-year (g, h) landmarks following type 2 diabetes diagnosis. Each model adds a successive layer of information: M1, intrinsic risk features only; M2, laboratory snapshots; M3, medication exposure; M4, longitudinal trajectories; M5, social determinants of health. All models were evaluated on an identical held-out test set under a fixed screening policy. The dashed horizontal line in each precision–recall panel indicates outcome prevalence at that landmark. AUC, area under the receiver operating characteristic curve; AUPRC, area under the precision–recall curve.

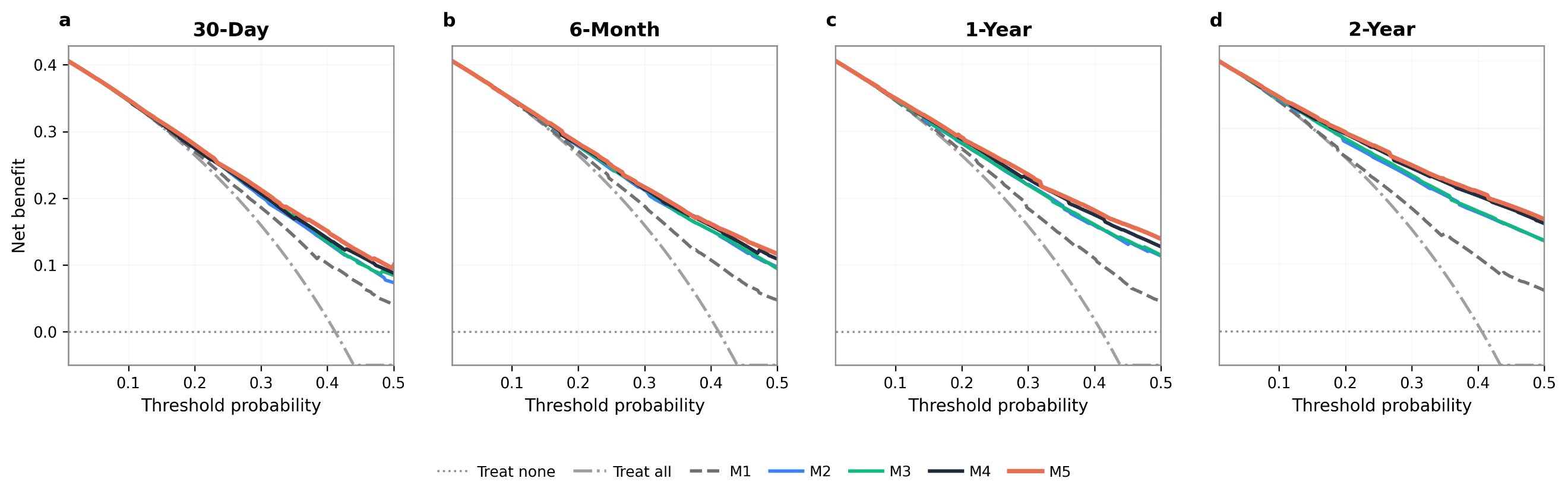

**Supplementary Figure S2. Decision curve analysis across prediction landmarks.**

Net benefit curves for models M1–M5, treat-all, and treat-none evaluated across a range of threshold probabilities at the 30-day (a), 6-month (b), 1-year (c), and 2-year (d) landmarks following type 2 diabetes diagnosis. Net benefit is defined as the true positive rate minus the false positive rate weighted by the odds of the threshold probability. The treat-all strategy assumes all individuals are screen-positive; the treat-none strategy assumes none are. Across all landmarks, models incorporating longitudinal trajectories (M4) and social determinants of health (M5) demonstrate consistently higher net benefit than earlier model layers and the treat-all strategy across the full range of clinically relevant thresholds. The incremental advantage of richer information layers over M1 and treat-all widens progressively from the 30-day to the 2-year landmark, reflecting the accumulation of longitudinal clinical signal over time.
